# Brain Functional Connectivity and Anatomical Features as Predictors of Cognitive Behavioral Therapy Outcome for Anxiety in Youths

**DOI:** 10.1101/2024.01.29.24301959

**Authors:** Andre Zugman, Grace V. Ringlein, Emily S. Finn, Krystal M. Lewis, Erin Berman, Wendy K. Silverman, Eli R. Lebowitz, Daniel S. Pine, Anderson M. Winkler

## Abstract

**Background:** Because pediatric anxiety disorders precede the onset of many other problems, successful prediction of response to the first-line treatment, cognitive-behavioral therapy (CBT), could have major impact. However, existing clinical models are weakly predictive. The current study evaluates whether structural and resting-state functional magnetic resonance imaging can predict post-CBT anxiety symptoms.

**Methods:** Two datasets were studied: (A) one consisted of n=54 subjects with an anxiety diagnosis, who received 12 weeks of CBT, and (B) one consisted of n=15 subjects treated for 8 weeks. Connectome Predictive Modeling (CPM) was used to predict treatment response, as assessed with the PARS; additionally we investigated models using anatomical features, instead of functional connectivity. The main analysis included network edges positively correlated with treatment outcome, and age, sex, and baseline anxiety severity as predictors. Results from alternative models and analyses also are presented. Model assessments utilized 1000 bootstraps, resulting in a 95% CI for *R*^2^, *r* and mean absolute error (MAE).

**Outcomes:** The main model showed a mean absolute error of approximately 3.5 (95%CI: [3.1-3.8]) points a *R*^2^ of 0.08 [−0.14 - 0.26] and *r* of 0.38 [0.24 – 0.511]. When testing this model in the left-out sample (B) the results were similar, with a MAE of 3.4 [2.8 – 4.7], *R*^2^ −0.65 [−2.29 – 0.16] and *r* of 0.4 [0.24 – 0.54]. The anatomical metrics showed a similar pattern, where models rendered overall low *R*^2^.

**Interpretation:** The analysis showed that models based on earlier promising results failed to predict clinical outcomes. Despite the small sample size, the current study does not support extensive use of CPM to predict outcome in pediatric anxiety.

## Introduction

As pediatric anxiety disorders precede the onset of most persistent adult emotional problems (Pine et al. 1998; Woodward and Fergusson 2001; Gregory et al. 2007; Nelemans et al. 2014), successful treatment could exert long-term impact. However, Cognitive Behavior Therapy (CBT), a first-line treatment, produces remission only in less than half of all cases (Ginsburg et al. 2011; Piacentini et al. 2014). Because CBT is time consuming, identifying reliable predictors of treatment outcome could markedly influence practice. Clinical features, such as comorbidity or severity only partially predict outcomes (Kunas et al. 2021). Magnetic resonance imaging (MRI) indices, in turn, may be able to predict outcomes beyond such clinical features; these measures are reliable, scalable, and already used in relatively large samples (Miller et al. 2016). The current study applies a predictive framework with resting-state functional connectivity (rsFC) and structural (sMRI) in medication-free children seeking treatment for anxiety disorders. Two samples are studied, each receiving CBT by trained experts, to support a three step approach. This begins with model building, followed by cross-validation in the first, larger sample. The approach ends with model testing in the smaller, held-out sample.

This study extends considerable research (Dubois and Adolphs 2016; Mueller et al. 2013) using rsFC to model an individual’s “connectome”, computed by correlating signals among network “nodes” (Sporns 2011). rsFC quantifies intrinsic network connections, which are stable and unique to the individual (Horien et al. 2019). Connectome Predictive Modeling (CPM) (Shen et al. 2017) generates clinical insights by correlating edgewise rsFC matrices with clinical measures and pooling associations in a second prediction stage. CPM can predict important constructs, such as intelligence (Finn et al. 2015; Greene et al. 2018; Gao et al. 2019), attention (Rosenberg et al. 2016; 2017), and anxiety (Wang et al. 2021; Ren et al. 2021). Although promising results exist, CPM is still understudied, as is rsFC for anxiety disorders more broadly (Zugman et al. 2023). Only two studies used rsFC analyzed with methods different from CPM, to predict treatment outcome in anxiety (Whitfield-Gabrieli et al. 2016; Ashar et al. 2021). Both studies examined adults, and the predictive model failed to replicate. No studies used CPM to predict treatment response in anxiety, and the available rsFC studies were small. Of note, while the sample size in the current study is also small, it is larger than either of the two past studies. Across the three studies, small sample sizes reflect the difficulty of delivering state-of-the-art treatment to medication-free subjects in the context of brain imaging investigations. Using such data, the primary goal of the current study is to predict CBT response using CPM in pediatric anxiety disorders.

The study’s secondary goal considers aspects of imaging reliability. On the one hand, structural MRI (sMRI) generates measures with greater reliability than rsFC. Hence, sMRI may have advantages in predicting treatment response. However, rsFC, while less reliable (Hedges et al. 2022; Noble, Scheinost, and Constable 2019), may identify a particular useful subset of stable features that relate more consistently than sMRI to some clinical measures (Mansour L et al. 2021). These measures, like anxiety symptoms, manifest as dimensional distributions, without sharp boundaries between patients and healthy people. Nevertheless, no study examines such measures using sMRI in a framework similar to CPM, let alone directly compares them with rsFC. We term the use of sMRI in this framework “Anatomical Predictive Modeling (APM)”, since the concept of connectome is not involved. Within the CPM framework, we compare the ability of sMRI and rsFC to predict treatment response.

Recent literature describes idiosyncratic rsFC patterns related to subject identity as akin to “fingerprints”. These patterns may predict variables of clinical interest (Finn et al. 2015; Amico and Goñi 2018; Byrge and Kennedy 2020; Lin et al. 2020). Recent research and commentary (Mantwill et al. 2022; Finn and Rosenberg 2021), however, suggest otherwise. Thus, to contribute to this ongoing discussion, a third objective of this study is to assess whether MRI features most unique to individuals predict response.

Thus, our work contributes to the literature in the following ways: (1) by providing an evaluation of the potential for use of CPM to predict CBT treatment response in anxious youth, (2) by formalizing an analogous procedure to CPM that uses brain morphology as predictors, as opposed to brain function, and (3) by evaluating whether the rsFC connections that most differentiate patients are predictive of treatment response.

## Methods

### Participants

Anxious youth and healthy volunteers were recruited through referral to participate in the study at the National Institute of Mental Health (NIMH), National Institutes of Health (NIH), Bethesda, MD, United States. They were enrolled as part of a clinical protocol (01-M-0192; Principal Investigator: Daniel S. Pine) for an ongoing clinical trial. Patients were considered for enrollment if they had a diagnosis of any DSM-5 anxiety disorder established by a licensed clinician using the KSADS (Kiddie Schedule for Affective Disorders and Schizophrenia). Exclusion criteria for all subjects were a history of psychotic disorder, bipolar disorder, developmental disorders, obsessive-compulsive disorder, post-traumatic stress disorder, substance use disorder, contraindication to MRI scan, use of medication, or an estimated IQ lower than 70 (as measured by the Wechsler Abbreviated Scale of Intelligence). HV also were excluded if they had any current psychiatric diagnoses. All parents and research participants provided written consent/assent in a protocol approved by the NIH Institutional Review Board (IRB). Symptom severity and treatment response was assessed using the Pediatric Anxiety Rating Scale (PARS) (The Research Units on Pediatric Psychopharmacology Anxiety Study Group 2002), the gold-standard clinician-administered assessment incorporating both child and parent report. The PARS was administered at four time points before, during (week 3 and week 8), and after treatment. The total score from the PARS is computed as recommended by the measures developers. Total PARS score ranges from 0-25, with a clinical cut-off of nine or higher indicating likely presence of an anxiety disorder. In addition to CBT administered by experts, all patients received either an active or sham version of attention-bias modification therapy (ABMT). To maximize sample sizes, groups were combined irrespective of randomization to either active or sham ABMT. CBT in this sample was delivered using a standardized protocol, consisting of 12 weekly sessions (Silverman et al. 2022; Silverman and Ginsburg 1998). The first three treatment sessions entail an introduction to CBT, psychoeducation, and self-monitoring/tracking. Starting at session four, participants complete in-session exposures and learn cognitive restructuring strategies and coping mechanisms (Lebowitz et al. 2019). For additional details, see (Haller et al. 2024).

The CPM analysis included only patients who had an available resting state fMRI scan collected up to 90 days before or 30 days after treatment initiation. As the primary objective was to assess treatment response, the main analysis excluded those who did not have available PARS at baseline or at 12 weeks. The baseline PARS came from either the screening visit or the third week of treatment and prior to the exposure-based portion of the treatment, and closer in time to the date of the MRI scan acquisition. Full sample description is provided in **Supplementary Table 1**. To facilitate comparison with CPM, the APM analysis considers the same individuals.

The fingerprinting analysis included only participants with at least two fMRI sessions within one year of each other. Aiming to maximize the number of subjects with two scans, fingerprinting analysis was conducted including both patients and healthy volunteers. It is important to note that subjects included in the primary data were allowed to be included in the fingerprinting analysis if they had a follow-up rsfMRI available. The sample characteristics are described in **Supplementary Table 2**. Fingerprinting with sMRI used the same individuals, to facilitate comparison between the two approaches.

To determine whether results obtained are replicable, we used a small sample of N = 15 individuals who participated in a previous 8 week clinical trial to study the effects of CBT on pediatric anxiety (White et al. 2017). This study was also under protocol 01-M-0192. It used a different resting state sequence, and CBT in this study followed the Coping Cat program (Podell et al. 2010). The eight sessions aim to develop skills to recognize signs of anxiety and anxious thoughts, relaxation techniques and coping. Again, we use only patients with resting state fMRI from up to 90 days before or 30 days after initiating treatment and who had available PARS at baseline and at the end of treatment (8 weeks). Demographic characteristics for this sample can be found in the Supplementary Material.

### Image acquisition

#### Dataset A

Scans were performed on two identical General Electric Signa Discovery MR750 3T scanners using a 32-channel head coil. Resting-state fMRI was acquired for 10 minutes (304 volumes) with a multi-echo planar sequence (TR: 2000 ms, TE: 14/28/42 ms, flip angle: 77°, FoV: 240×240 mm, matrix size: 64×64, 34 axial interleaved slices, bandwidth: 7812.5 Hz/Pixel, voxel size: 3.75×3.75×3.80 mm). Participants were instructed to keep their eyes open and focus on a white fixation cross in the middle of a black screen. A T1-weighted MPRAGE sequence was acquired in the same scanning session (TR: 7.66 ms, TE: 3.476 ms; TI: 900 ms^1^, FoV: 256×256 mm, flip angle: 7°, voxel size: 1×1×1 mm). Patients were allowed to watch a movie of their choice during the non-functional part of the scanning session. Dataset A was used for fingerprinting, and for CPM and APM with cross-validation.

#### Dataset B

For the replication analysis, a dataset collected in one of same scanners was used, although these images used a different echo-planar sequence for the fMRI (TR: 2000 ms, TE: 30 ms (single echo), flip angle: 90°, FoV: 192×192 mm, matrix size: 64×64, 36 axial interleaved slices, bandwidth: 7812.5 Hz/Pixel, voxel size: 3.0×3.0×4.0 mm), that was also shorter (6 minutes, 180 volumes), as well as for the corresponding T1-weighted MPRAGE (TR: 7.82 ms, TE: 3.02 ms, TI: 725 ms, FoV: 256 × 256 mm, flip angle: 6°, voxel size: 0.86×0.86×1.20 mm). Patients were, likewise, allowed to watch a movie of their choice acquisition of the non-functional scans. Dataset B was used for external validation of CPM and APM.

### Image processing

For both datasets, images were preprocessed identically using fMRIPrep (v. 21.0.0) (Esteban et al. 2019). Additional QC metrics were obtained with MRIQC (v. 0.16.1) (Esteban et al. 2017). All relevant outputs were visually inspected. Subjects were excluded for excessive motion if they had a frame displacement mean of greater than 0.25 mm or more than 50% of frames with frame displacement greater than 0.25 mm, and appeared to be low quality based on visual inspection of the fMRIPrep outputs. Of the 66 patients in Dataset A available for the CPM and APM analyses, n = 12 were excluded based on these criteria, leaving n = 54 subjects for the primary analysis. Of the 87 (patients = 57) subjects available for the fingerprinting analyses, n = 21 (patients = 15) were excluded based on these criteria, leaving n = 66 (patients = 42). Of the 31 patients in Dataset B available for CPM and APM, 16 were excluded based on these criteria, leaving n = 15 patients.

Details of motion and demographics in each sample are in **Tables 1 and 2**, and in **Supplementary Tables 1 and 2**; demographics for the excluded participants can be found in **Supplementary Tables 3, 4, and 5**. The excluded group for the fingerprinting analysis were significantly lower in baseline age (p = 0.02) and follow-up age (p = 0.03) than the included groups but were not significantly different in age of scan in the in Dataset A of the CPM analysis (Dataset A: p = 0.32, Dataset B: p < 0.01). Removal of motion artifacts was performed by regressing out ICA-AROMA noise components (Pruim et al. 2015), global signal, white matter signal, and CSF signal from the images aligned to MNI space. We use global signal regression (GSR) given that it has been shown to increase the correlation between resting state functional connectivity and behavioral measures (Li et al. 2019). Results without GSR are shown in the Supplementary Material.

**Table 1:**
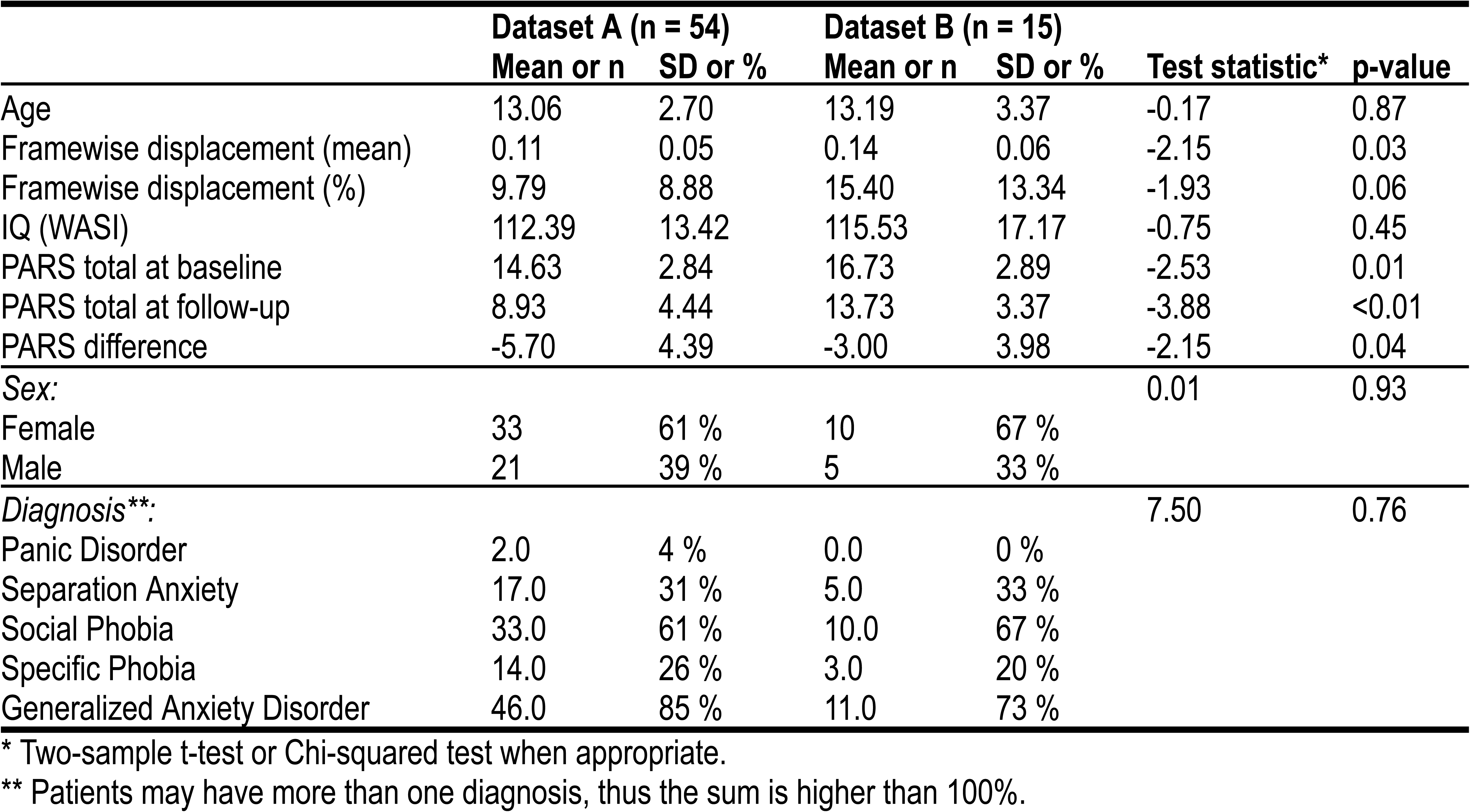
Descriptive statistics for Datasets A and B, as used for the CPM and APM analyses. Additional sample details can be found in the **Supplementary Material**.

**Table 2:**
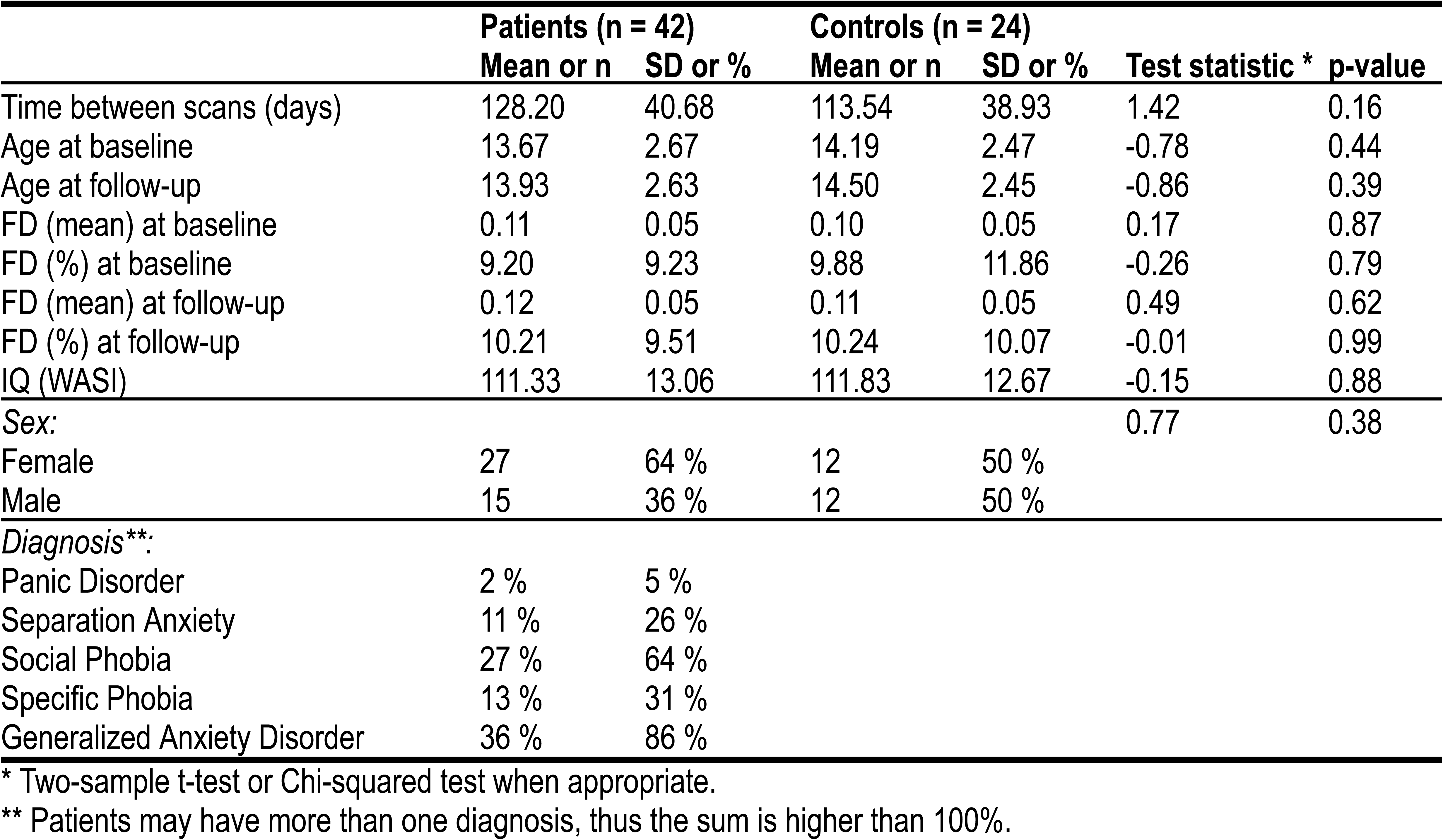
Descriptive statistics for the fingerprinting sample (from Dataset A). Additional sample details can be found in the **Supplementary Material**.

### Functional connectivity

The processed images were parcellated, in surface space, according to the Schaefer atlas with 200 surface-based ROIs (Schaefer et al. 2018) (100 per hemisphere), grouped into 17 networks (Yeo et al. 2011) and additional 16 FreeSurfer (Fischl 2012) ROIs for subcortical structures (8 for each hemisphere). A correlation matrix representing the rsFC between every pair of regions was computed for each subject and each session using Pearson’s correlation (*r*), and converted to a z-score using Fisher’s transformation (*z* = arctanh(*r*)), resulting in 23,220 unique edges that were used for fingerprinting and for CPM. Two variants of rsFC were considered: simple correlations, as well as partial correlations, whereby the timecourses of all other regions are regressed out from the timecourses of every pair of nodes whose correlation is being computed.

### Connectome Predictive Modeling

CPM analysis followed the method outlined in (Shen et al. 2017). Before conducting CPM, we verified that motion (as assessed via average framewise displacement) would not be a good predictor of the PARS score (Dataset A: r = −0.2030, p = 0.1380; Dataset B: r = 0.0877, p = 0.7544; two-tailed p-values assessed with 10,000 permutations). The rsFC matrices for every subject were tested for their association with PARS at the end of treatment; significantly (p < 0.01) associated edges were selected, and their rsFC r-to-z values summed. These sums were used as independent variables in a second linear regression.

#### Cross validation

Regression coefficients from the second model were used to predict, in a leave-one-out cross-validation loop, the PARS score of a subject unseen in the previous steps. This analysis used the patients from Dataset A.

#### External validation

Regression coefficients from the second model with Dataset A were used to predict the PARS score of all patients in Dataset B.

Selection of significant edges in the initial step of CPM can consider edges that are positively correlated with PARS, negatively correlated, or both; edges can also be selected using other criteria. We departed from (Shen et al. 2017) in two aspects: (1) we investigated the inclusion of age and sex in the second regression model as predictors of interest and, separately, as nuisance, as well as without any such additional regressors as in the original publication; and (2) in addition to investigating the performance of using only positively correlated, negatively correlated, and both sets of edges, we also investigated the performance of CPM when using the most discriminative edges identified using fingerprinting; details of these two departures from the original method are provided below, and results from these various models are presented in the Supplementary Material. Model performance was assessed using the mean absolute error (MAE), the simple correlation coefficient (*r*) between observed (*y_i_*) and predicted values 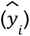 and a version of the coefficient of determination (*R*^2^) that is suitable for cross-validation and is computed as (Kvålseth 1985):

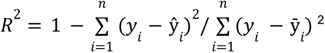

Note that *R*^2^ does not correspond to the square of the correlation coefficient *r*; for commentary on the merits of each metric, see (Poldrack, Huckins, and Varoquaux 2020; Chicco, Warrens, and Jurman 2021). Confidence intervals (95%) were computed for these three quantities using 1000 bootstraps (Davison and Hinkley 1997).

### Anatomical Predictive Modeling

We investigated how replacing rsFC in CPM with measurements of brain morphology, which we termed APM, would impact predictions. Surface-based representations of the brain were obtained with FreeSurfer 6.0.1, as part of fMRIprep processing, and resampled into the “fsaverage5” space (a brain mesh with the same topology of a geodesic sphere produced by 5 recursive subdivisions of an icosahedron). The fsaverage5 contains 20,484 vertices spanning both hemispheres; we removed those with constant variance, thus masking out non-cortical regions, to a total of 18,742 vertices used for analysis (compare to 23,220 unique edges used for analysis in the rsFC-based models). We investigated 5 different cortical morphometric measurements (area, thickness, curvature, sulcal depth, and gray/white matter contrast), and two levels of smoothing (FWHM = 0 and 15 mm).

### Nuisance variables

Prediction may make use of other variables, such as age and sex, or consider these as nuisance. In the former case, they are included as additional regressors in both stages of CPM and, subsequently, as additional predictors (Rao, Monteiro, and Mourao-Miranda 2017). In the latter case, these variables are likewise included in the first regression model of CPM (that identifies edges), whereas in the second regression, both data and model are residualized with respect to these variables in the training set; the estimated regression coefficients from the training set are then to residualize also the test set (Snoek, Miletić, and Scholte 2019); prediction uses then residualized variables, with coefficients of variables of interest and of no interest estimated from the training set. Nuisance effects can be added back to the predicted values to ensure results are compatible with the quantities of interest. If the data used for testing contains a substantial number of subjects, an improved model consists of residualizing the test set using estimated nuisance effects from the test set itself, as opposed to from the training set, thus reducing the risk of covariate shift (Rao, Monteiro, and Mourao-Miranda 2017). We investigated models without nuisance variables, as well as with age and sex (and scanner where appropriate). For the cross-validation case, in which it is not possible to estimate nuisance effects from test samples (in a leave-one out cross-validation, the test set has only one subject), we used the regression coefficients for nuisance variables obtained from the training set (Snoek, Miletić, and Scholte 2019), whereas for external validation, nuisance effects were estimated directly from the test set.

### Fingerprinting

Fingerprinting analysis followed the methods outlined in (Finn et al. 2015). Each subject had two resting state scans, collected on average 123 days apart. The rsFC matrix was unwrapped into a vector (only the upper or lower triangular part is needed since the rsFC matrix is symmetric around the main diagonal), then the Pearson’s correlation coefficient between every baseline rsFC of every subject with every follow-up rsFC was computed, providing an index of similarity. Subject identification was considered successful if, for every baseline rsFC matrix, the most similar follow-up rsFC matrix belongs to the same subject. As the most similar follow up is allowed to be repeated (i.e., prediction is done with replacement), the p-value for the number of correct identifications (*k*) can be computed analytically (without the need for permutations) using the cumulative distribution function of the binomial distribution with parameters *n* = number of subjects, *p* = 1/*n* and location *k* = 1.

Correlations can be interpreted as the dot product of vectors normalized to unit variance (Rodgers and Nicewander 1988). This provides an indicator of the contribution of each edge to the final correlation. Let the correlation be expressed as (Finn et al. 2015):

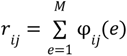

where *M* is the number of edges, 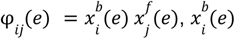 is the normalized value of the rsFC at edge *e* for subject *i* at baseline, and 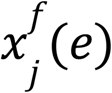 is the normalized rsFC at the same edge for subject *j* at follow-up. If *i* = *j*, *r_ii_* is the correlation between a subject’s own baseline and follow-up rsFC matrices. The quantity *φ* is interesting because, if *φ_ii_*(*e*) ≥ *φ_ij_*(*e*) and *φ_ii_*(*e*) ≥ *φ_ji_*(*e*), then edge *e* contributes to the identification of the subject’s rsFC at the other time point. An estimator of the probability *P_i_*(*e*) that an edge makes such contribution by chance is given by:

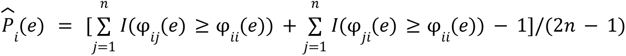

where I(·) is the indicator (Kronecker) function, which evaluates as 1 if the condition inside parentheses is true, 0 otherwise, and *n* is the number of subjects. Note that the above formulation is different than the one originally proposed by (Finn et al. 2015); edges that are highly predictive can have 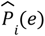 as low as 1/(2*n* − 1), as opposed to zero; edges that are not predictive can have 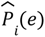 as high as 1. A global estimate of the differential power (DP) of a given edge for subject identification can be computed as:

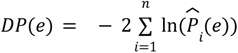

where the 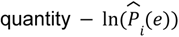 follows an exponential distribution with rate parameter 1 if the true (unknown) *P_i_*(*e*) follows a uniform distribution. The constant 2 adjusts that rate to 1/2. An exponential distribution with rate parameter 1/2 is a Chi-squared distribution with 2 degrees of freedom; the sum of *n* random variables following this distribution also follows a Chi-squared distribution, now with 2*n* degrees of freedom. Thus, the hypothesis that an edge is more informative than could be expected by chance can be tested. This formulation also allows the selection of edges (e.g., for later analyses, such as in CPM) using a threshold based on the probability distribution under the null hypothesis of chance differential power. Note that above formulation of *DP*(*e*) is also different from the original work by (Finn et al. 2015).

#### Anatomical Fingerprinting

Following (Mansour L et al. 2021), we also investigate fingerprinting using measures of cortical morphology: area, thickness, curvature, sulcal depth, and gray/white matter contrast, as opposed to only unwrapped rsFC matrices. Fingerprinting methods are otherwise the same as for rsFC data. Surface-based cortical measurements were as with APM.

## Results

### Prediction of anxiety scores using CPM

We report main results for CPM using GSR, full (not partial) rsFC, positive edges without weighting, and both a model in which age, sex, and baseline PARS are used as predictors, as well as a model in which data are residualized in relation to these variables. These results are emphasized since they led to generally superior accuracy across multiple analyses. CPM was not able to predict post-treatment 12-week PARS scores at a level that exceeded chance (**Table 3**). While using the simple correlation coefficient might give the impression of statistically robustness, the magnitude of relations between predicted and expected scores was only moderate, with a mean absolute error of approximately 3.5 points (PARS scores range between 0 and 25). This mean absolute error does approach a level that would be clinically useful (Walkup et al. 2001; 2008), but the correlation between predicted and observed PARS did not exceed 0.4. Additionally, no model showed a high *R*^2^, and the R^2^ 95% CI indicated that no model was better than chance. Overall, the low *R*^2^ indicates that the models fail to predict above the mean of the target value. A scatter plot showing observed and predicted values for one of these models appears in **Figure 1, upper left panel**.

**Figure 1:**
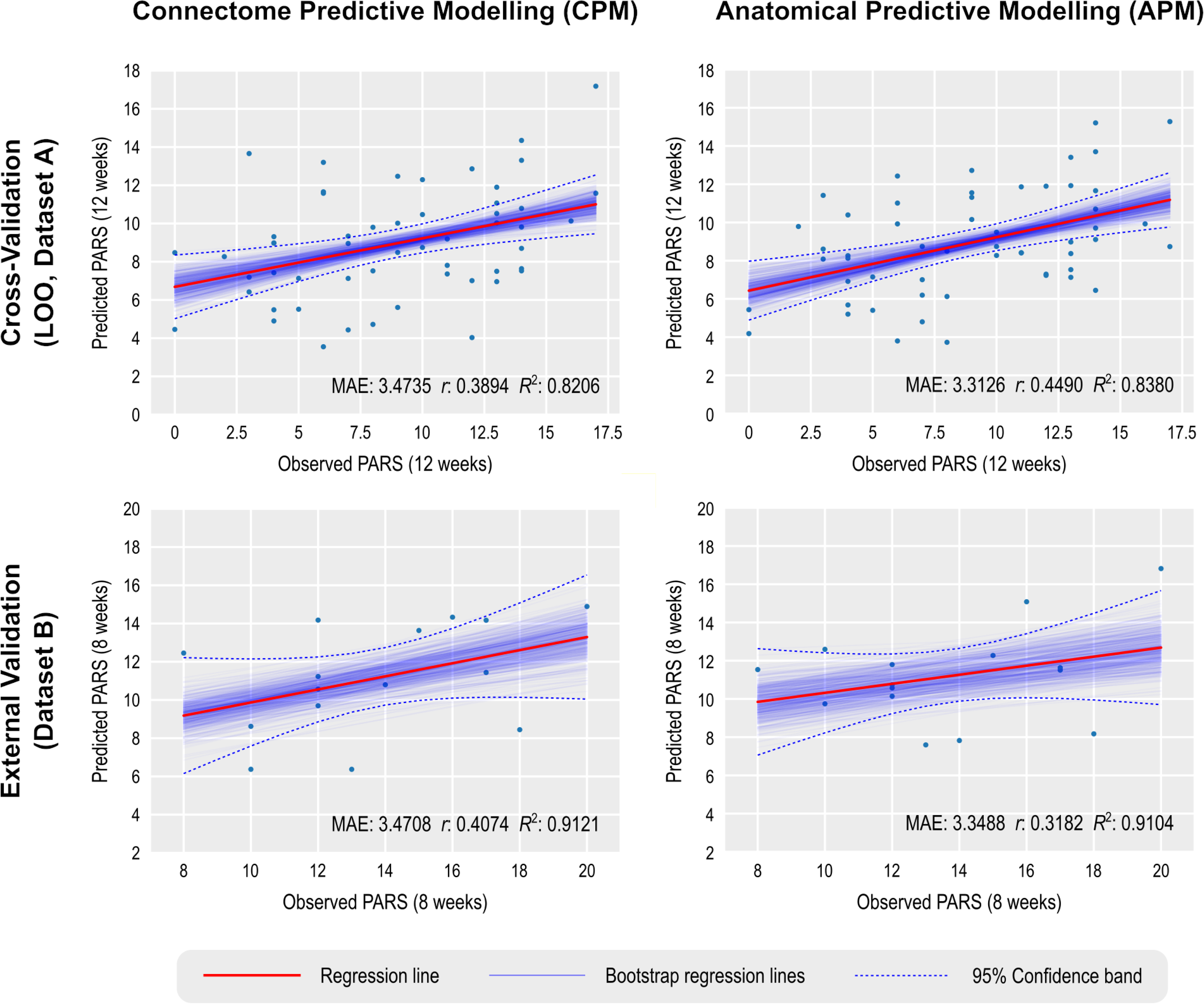
Prediction of anxiety scores using CPM and APM; APM used gray/white matter contrast. The main regression line (red) is based on the observed and predicted values (represented by the dots). The bootstrap regression lines (faint blue) are based on the bootstrapped predictions used to construct the 95% confidence intervals shown in Tables 3 and 4 (to avoid clutter, only 500 out of 1000 lines are shown in each panel). The 95% confidence bands were computed parametrically in relation to the main regression line, and is presented merely as an additional reference. Observe that the slopes of the bootstrapped lines in the external validation are less variable, which is expected given the larger number of observations that are predicted in a single step (15 in this case) versus the single prediction in each step of the leave one out (LOO) cross-validation.

**Table 3:**
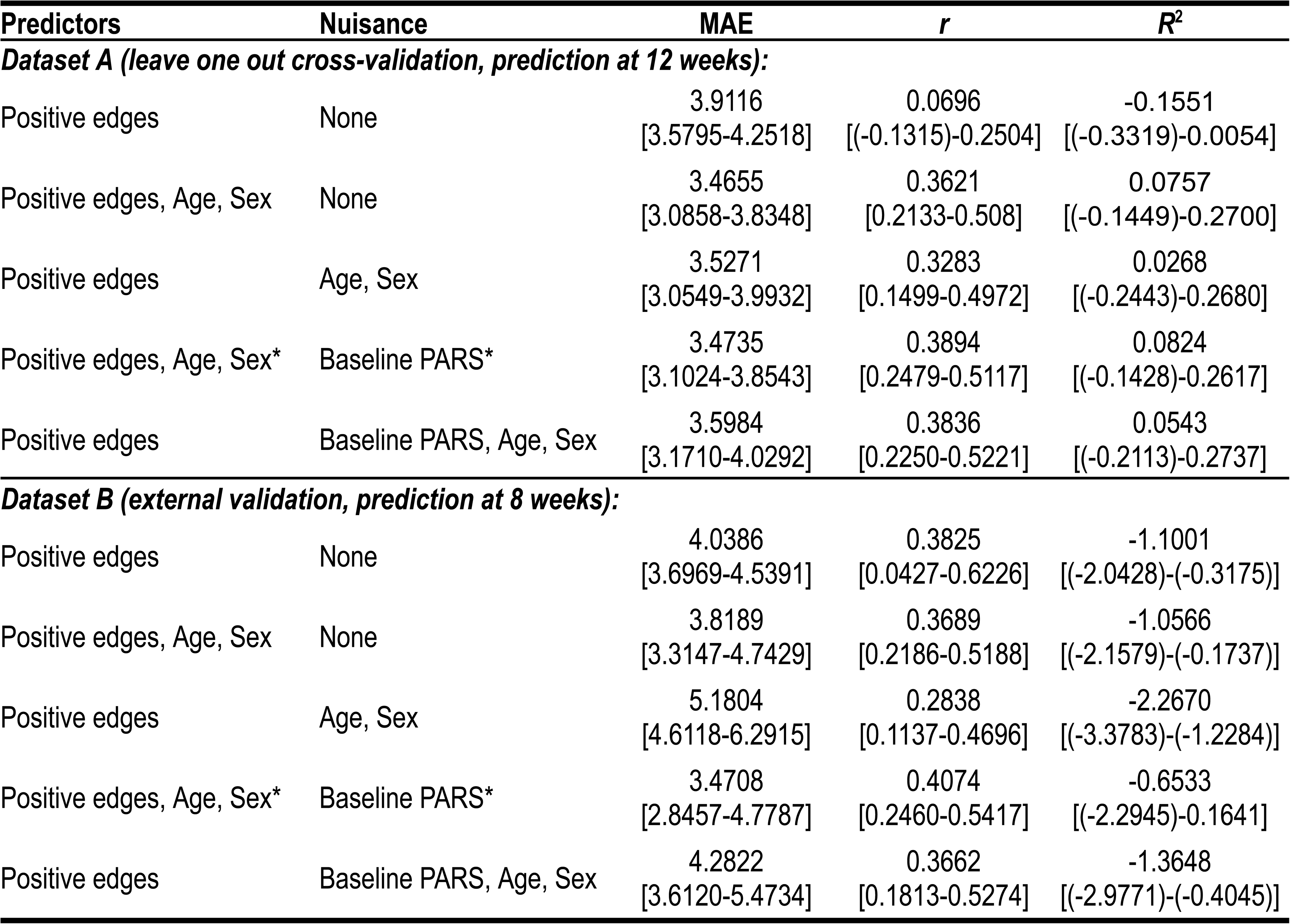
Mean absolute error (MAE) of CPM-predicted vs. observed PARS at 12 weeks, using Dataset A for training and leave one out cross-validation, and Dataset B for external validation. The corresponding correlation (*r*) and coefficient of determination (*R*^2^) are also shown. Confidence intervals (95%), based on 1000 bootstraps, are between brackets. A scatter plot for the model marked with an asterisk (*) is in **Figure 1 (left panels)**.

| Predictors | Nuisance | MAE | $r$ | $R^2$ |
| --- | --- | --- | --- | --- |
| **Dataset A (leave one out cross-validation, prediction at 12 weeks):** |  |  |  |  |
| Positive edges | None | 3.9116<br>[3.5795-4.2518] | 0.0696<br>[(-0.1315)-0.2504] | -0.1551<br>[(-0.3319)-0.0054] |
| Positive edges, Age, Sex | None | 3.4655<br>[3.0858-3.8348] | 0.3621<br>[0.2133-0.508] | 0.0757<br>[(-0.1449)-0.2700] |
| Positive edges | Age, Sex | 3.5271<br>[3.0549-3.9932] | 0.3283<br>[0.1499-0.4972] | 0.0268<br>[(-0.2443)-0.2680] |
| Positive edges, Age, Sex* | Baseline PARS* | 3.4735<br>[3.1024-3.8543] | 0.3894<br>[0.2479-0.5117] | 0.0824<br>[(-0.1428)-0.2617] |
| Positive edges | Baseline PARS, Age, Sex | 3.5984<br>[3.1710-4.0292] | 0.3836<br>[0.2250-0.5221] | 0.0543<br>[(-0.2113)-0.2737] |
| **Dataset B (external validation, prediction at 8 weeks):** |  |  |  |  |
| Positive edges | None | 4.0386<br>[3.6969-4.5391] | 0.3825<br>[0.0427-0.6226] | -1.1001<br>[(-2.0428)-(-0.3175)] |
| Positive edges, Age, Sex | None | 3.8189<br>[3.3147-4.7429] | 0.3689<br>[0.2186-0.5188] | -1.0566<br>[(-2.1579)-(-0.1737)] |
| Positive edges | Age, Sex | 5.1804<br>[4.6118-6.2915] | 0.2838<br>[0.1137-0.4696] | -2.2670<br>[(-3.3783)-(-1.2284)] |
| Positive edges, Age, Sex* | Baseline PARS* | 3.4708<br>[2.8457-4.7787] | 0.4074<br>[0.2460-0.5417] | -0.6533<br>[(-2.2945)-0.1641] |
| Positive edges | Baseline PARS, Age, Sex | 4.2822<br>[3.6120-5.4734] | 0.3662<br>[0.1813-0.5274] | -1.3648<br>[(-2.9771)-(-0.4045)] |

Of note, as expected, results were weaker in some analyses appearing in **Supplemental Material**, where a larger set of results, with varying processing choices, are provided. This included analyses predicting change scores and analyses using the smaller Dataset B to build the model, which was then tested in the larger Dataset A **(Figure 1, lower left panel)**.

### Prediction of anxiety scores using APM

For APM, the models that produced generally better models were those that used the gray/white matter contrast, without smoothing, and that selected both positive and negative vertices in the first regression of APM. Moreover, this set of APM models also tended to produce higher correlations between predicted and observed PARS scores than CPM, with correlations above 0.4. However, as with CPM, the results show no model produced a strong *R*^2^. A summary of the results is presented in **Table 4**, and a scatter plot showing observed and predicted values for one of these models appears in **Figure 1, upper right panel**.

**Table 4:** Mean absolute error (MAE) of APM-predicted (with gray/white contrast) vs. observed PARS, using Dataset A for training and leave one out cross-validation, and Dataset B for external validation. The corresponding correlation (*r*) and coefficient of determination (*R*^2^) are also shown. Confidence intervals (95%), based on 1000 bootstraps, are between brackets. A scatter plot for the model marked with an asterisk (*) is in **Figure 1 (right panels)**.

| Predictors | Nuisance | MAE | $r$ | $R^2$ |
| --- | --- | --- | --- | --- |
| **Dataset A (leave one out cross-validation, prediction at 12 weeks):** |  |  |  |  |
| Both pos/neg vtx | None | 3.3416<br>[3.0283-3.6602] | 0.4074<br>[0.2052-0.5714] | 0.1539<br>[(-0.0183)-0.3104] |
| Both pos/neg vtx, Age, Sex | None | 3.2228<br>[2.8900-3.5997] | 0.4242<br>[0.2846-0.5386] | 0.1515<br>[(-0.0648)-0.3301] |
| Both pos/neg vtx | Age, Sex | 2.9331<br>[2.1735-3.7533] | 0.5421<br>[0.3514-0.6926] | 0.2823<br>[(-0.2671)-0.7636] |
| Both pos/neg vtx, Age, Sex* | Baseline PARS* | 3.3126<br>[2.9656-3.6979] | 0.4490<br>[0.3108-0.5619] | 0.1714<br>[(-0.0462)-0.3692] |
| Both pos/neg vtx | Baseline PARS, Age, Sex | 3.9326<br>[3.1805-4.7093] | 0.4579<br>[0.2765-0.6103] | -0.1029<br>[(-0.6948)-0.3560] |
| **Dataset B (external validation, prediction at 8 weeks):** |  |  |  |  |
| Both pos/neg vtx | None | 3.3873<br>[2.9435-4.19478] | 0.2376<br>[(-0.2450)-0.6356] | -0.7177<br>[(-1.9185)-0.2394] |
| Both pos/neg vtx, Age, Sex | None | 4.5185<br>[3.8783- 5.7451] | 0.1709<br>[0.0177-0.3664] | -1.7479<br>[(-3.0675)-(-0.6599)] |
| Both pos/neg vtx | Age, Sex | 5.1874<br>[4.5683-6.3311] | 0.2283<br>[0.0544-0.4180] | -2.4419<br>[-3.6649-(-1.4652)] |
| Both pos/neg vtx, Age, Sex* | Baseline PARS* | 3.3488<br>[2.6322-4.7357] | 0.3182<br>[0.1532- 0.4607] | -0.6847<br>[(-2.197)-0.2572] |
| Both pos/neg vtx | Baseline PARS, Age, Sex | 4.2541<br>[3.5672-5.4248] | 0.3517<br>[0.1782-0.5139] | -1.4368<br>[-2.8349-(-0.5182)] |

Unlike for the cross-validation results, models for CPM generally produced stronger results for external validation than models for APM. Moreover, whereas results for CPM appeared generally comparable across cross-validation and external validation, for APM, external validation for Dataset B produced indices of accuracy that were generally lower than for cross-validation. **Figure 1, lower right panel** shows the corresponding scatter plot for observed and predicted values for Dataset B. An extended set of results for APM with cortical thickness, surface area, curvature, and sulcal depth, with and without smoothing, are provided in the **Supplementary Material**.

### Localization of predictive edges and vertices

For both CPM and APM, the predictive elements – edges or vertices, respectively – identified in the first regression manifested topographies that appeared widely distributed throughout the brain. These topographies did not manifest patterns comparable to networks identified in other research highlighting network-specific functions. We focus on the models that included age, sex, and baseline PARS as nuisance; these are highlighted with an asterisk (*) in **Tables 3 and 4**. **Figure 2** shows the edges most frequently identified in the leave one out cross-validation using Dataset A with CPM; **Figure 3** provides a similar depiction for APM, using gray/white matter contrast. For both CPM and APM, the number of elements found as significant in the first stage of the respective predictive model was relatively small, about two orders of magnitude smaller than the number of edges or vertices available for a given model.

**Figure 2:**
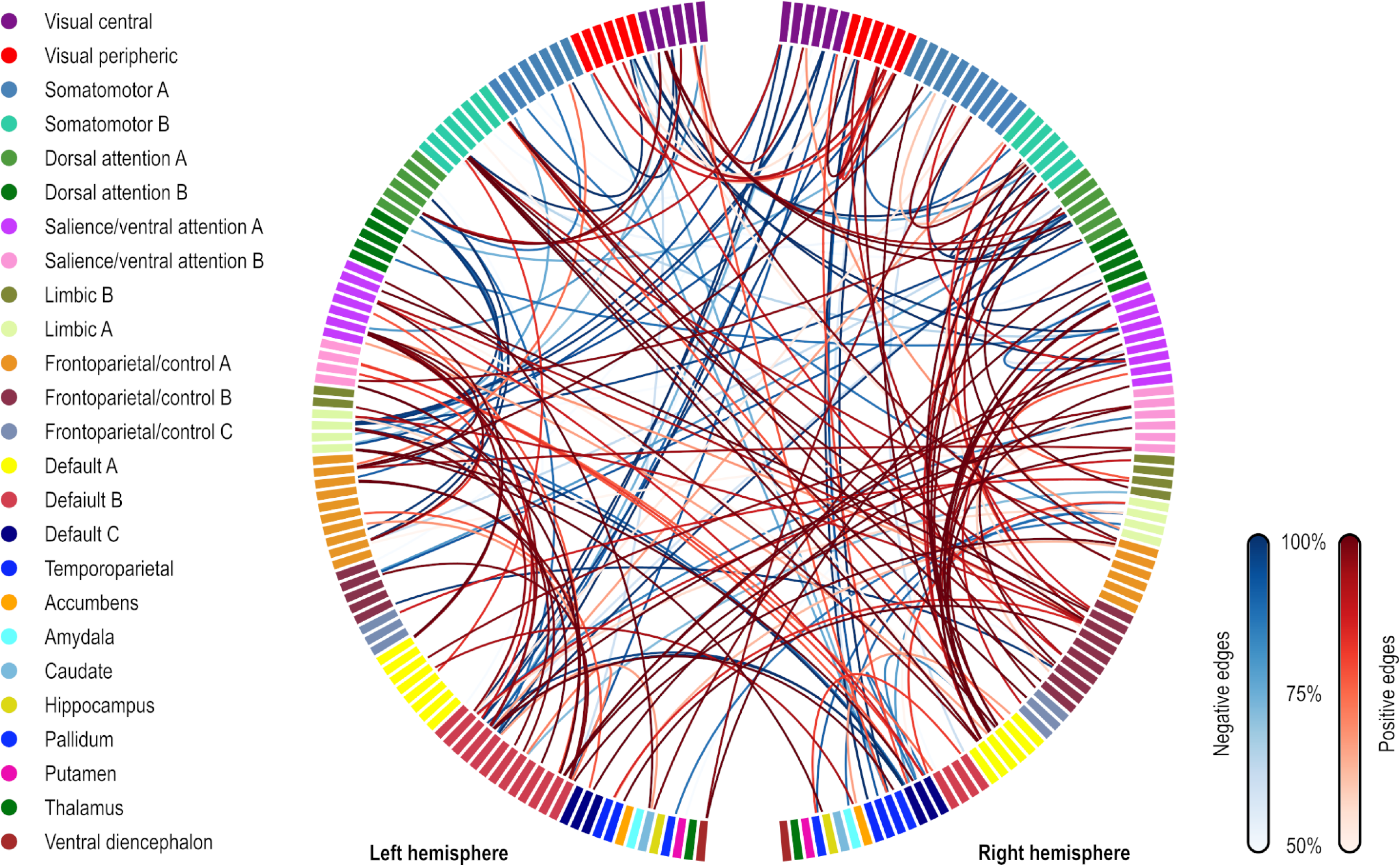
Edges most frequently identified as positively (red) or negatively (blue) associated with the PARS score at 12 weeks in Dataset A, as found in the first stage of CPM. The frequency refers to the number of iterations of the leave one out cross-validation in which a significant association was found; edges found in at least 50% of the iterations are shown (128 positive, 73 negative, out of 23,220 edges). The connections shown are for the model marked with an asterisk (*) in **Table 3** (only the positive edges were used in the second stage of CPM; the negative edges are depicted for completeness). Named networks are those identified by (Yeo et al. 2011); the set of nodes also includes 8 subcortical regions. Note that despite the seemingly large number of connections, only a small fraction of the total number of edges is used, in a pattern mostly diffuse and unstructured.

**Figure 3:**
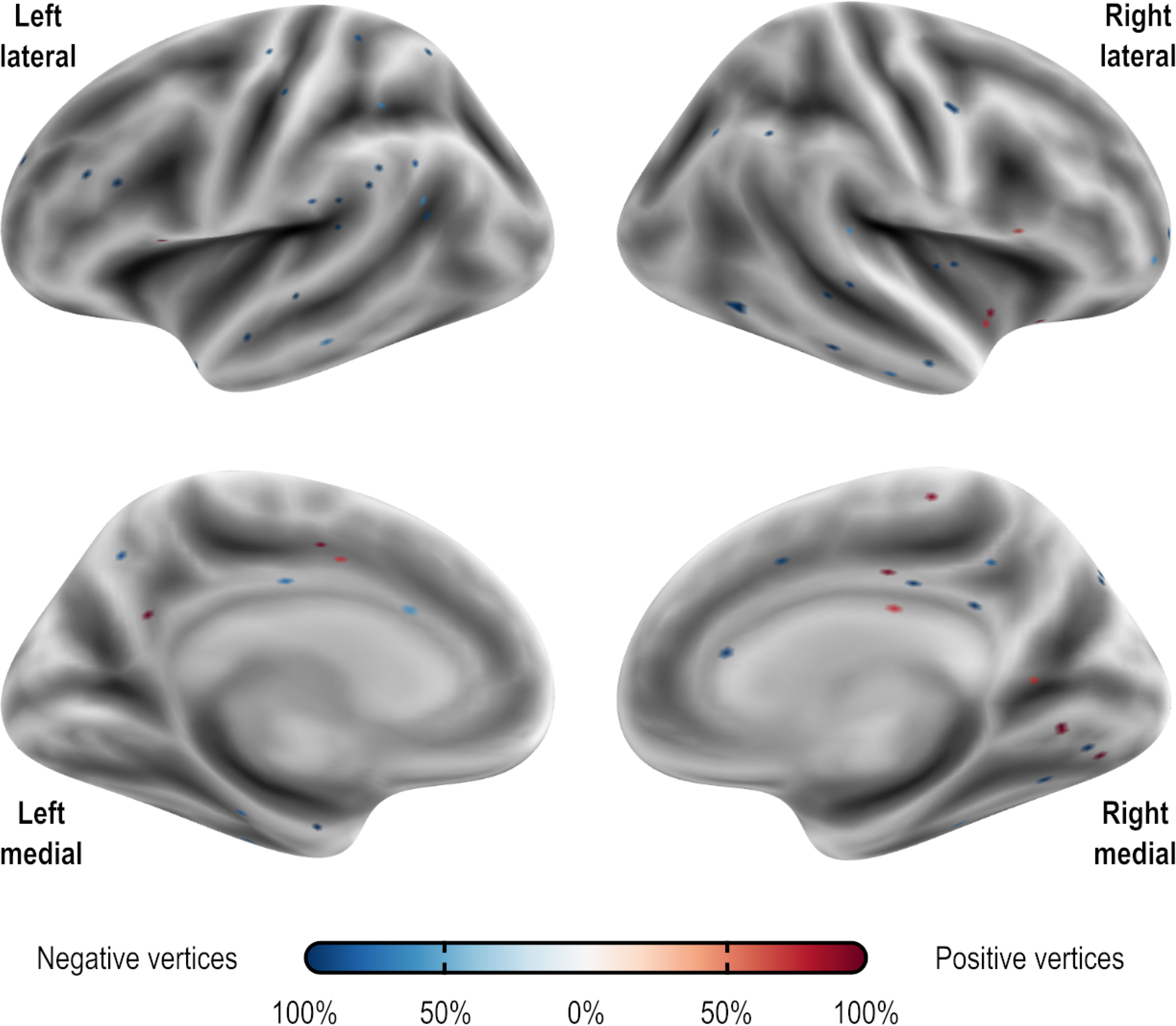
Vertices most frequently identified as positively (red) or negatively (blue) associated with the PARS score at 12 weeks in Dataset A. Observe that the pattern is mostly scattered, with isolated vertices (representing tiny regions) diffusely spread throughout the cortex. These results are as found in the first stage of APM using gray/white matter contrast. The percentage refers to the number of iterations of the leave one out cross-validation in which a significant association was found over all iterations; vertices found in at least 50% of the iterations are shown (16 positive and 55 negative, out of 18,742 vertices). The vertices shown are for the model marked with an asterisk (*) in **Table 4**.

### Uniqueness and its predictive value

Fingerprinting using rsFC features and sMRI features led to strong accuracy for subject identification. Using rsFC features from the baseline scan, the correct follow-up scan was correctly identified for 53 of 66 subjects (80.3%, p = 6.2×10^−84^), whereas doing the reverse produced correct identifications for 56 of 66 (84.9%, p = 2.3×10^−91^); these results are based on full (not partial) correlations, and with GSR. Anatomical fingerprinting led to even higher rates of correct identification, with near 100% success rate for most of the measurements studied (cortical area, thickness, curvature, sulcal depth, and gray/white matter contrast). Differential power for edges and for the gray/white matter contrast are shown in **Figure 4**, as p-values and in logarithmic scale, to allow scale comparisons and improved visual contrast; **Figure 5** shows DP for the other anatomical measurements. DP was found substantially higher for every anatomical measurement studied compared to connectivity measurements: while DP for edges were found generally weak and scattered, for gray/white contrast, DP was found stronger and with well-defined locations, covering mostly parietal cortex, precuneus, inferior temporal lobe, and caudal portions of the frontal lobe before reaching the precentral gyrus, and preserving central sulcus, pre- and postcentral gyri, insula, and cuneus, all of which are regions of known lower variability among individuals. The relation between DP in different modalities can be seen in suppl. **Figure 1**. There is little relation between the DP between structural measurements and rsFC.

**Figure 4:**
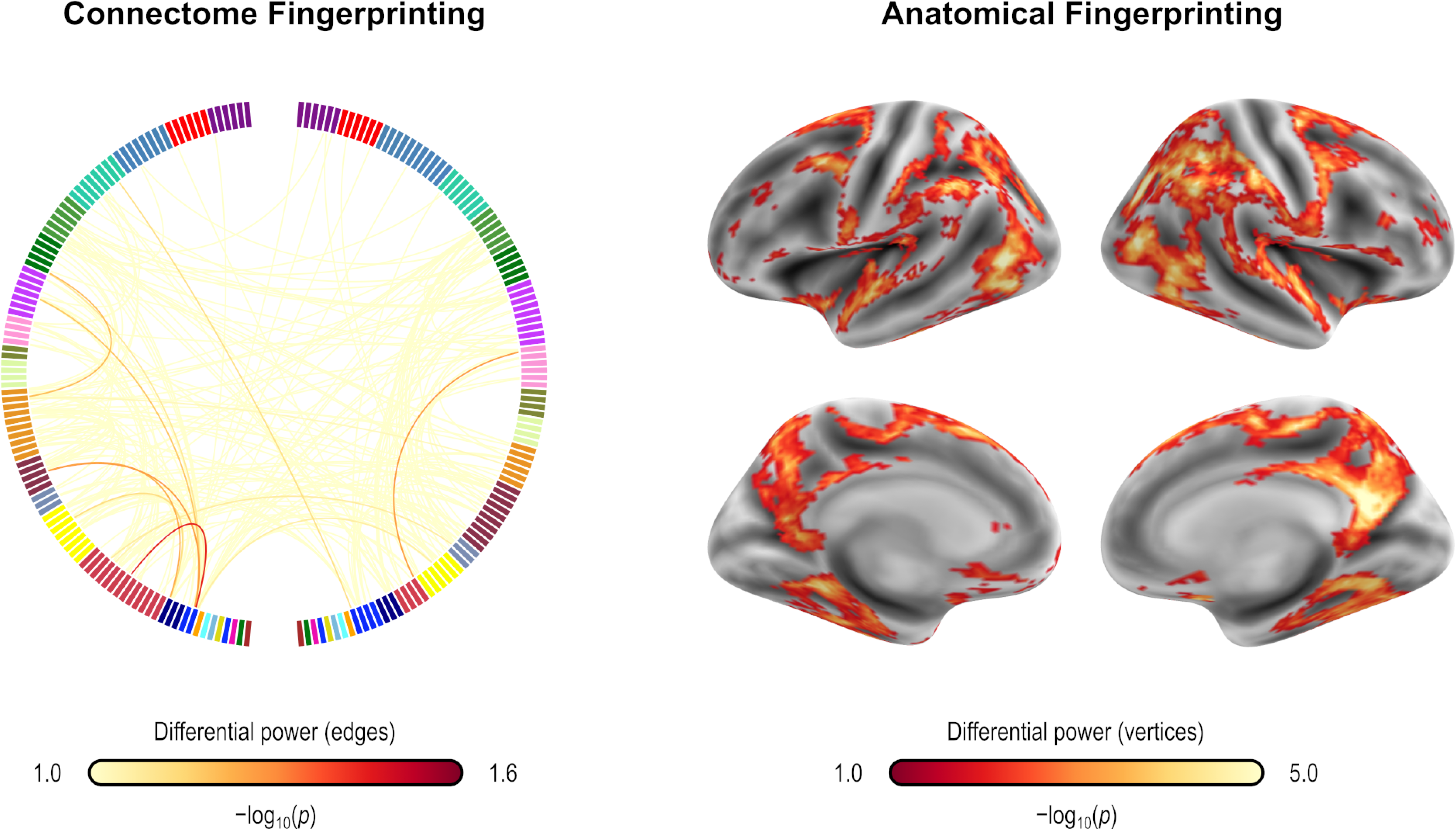
Differential power (DP) for edges using connectome fingerprinting (*left*), and for vertices using anatomical fingerprinting with gray/white matter contrast (*right*), in logarithmic scale based on their p-values (that is, −log_10_(*p*), where *p* is the p-value for DP, thus allowing scales to be comparable). Network names for the left panel are the same as for Figure 2, and name views are the same as for Figure 3. While anatomical fingerprinting without smoothing was slightly more accurate, the smoothed version includes the same regions and is more informative, hence it is the one shown. Higher values for the differential power indicate features that are more unique. DP is much higher for anatomical measurements than for connectivity measurements (note the different color scales); DP for connectivity features (edges) is generally weak and scattered, whereas for gray/white contrast (vertexwise), DP is stronger and with better defined localization. DP for cortical area, thickness, curvature, and sulcal depth are shown in Figure 5.

**Figure 5:**
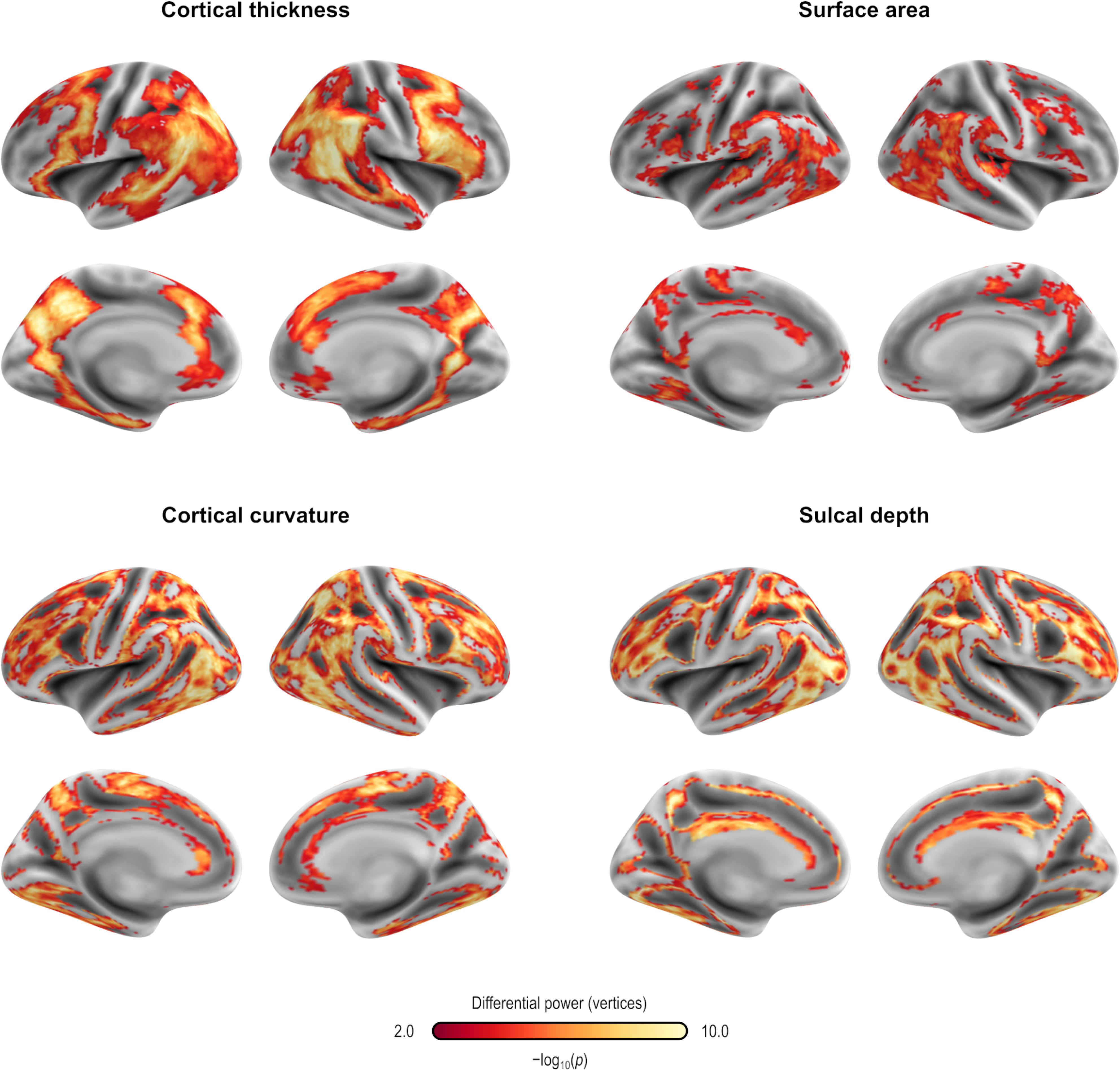
Differential power (DP) for vertices using anatomical fingerprinting with cortical thickness, cortical surface area, cortical curvature, and sulcal depth, in logarithmic scale based on their p-values (that is, −log_10_(*p*), where *p* is the p-value for DP, thus allowing scales to be comparable). While anatomical fingerprinting without smoothing was slightly more accurate, the smoothed version includes the same regions and is more informative, hence it is the one shown. Higher values for the differential power indicate features that are more unique. As with gray/white contrast, DP for other anatomical measurements was higher for non-primary, associative areas, where anatomical variability is also higher.

The edges or vertices with higher DP derived from fingerprinting, that is, those more “unique”, yielded slightly lower correlation to PARS scores compared to those found by model fitting in the first stage of CPM/APM. There was no overlap between the edges selected by using fingerprinting, compared to the edges found in the first stage of the CPM approach; the same was observed for APM.

## Discussion

This work applied CPM to predict response to CBT in pediatric anxiety disorders. The study used expert clinicians and a gold-standard measure of treatment outcome, in medication-free subjects recruited using criteria from past large-scale randomized controlled trials of pediatric anxiety disorders, i.e., RUPP 2001 (Walkup et al. 2001) and CAMS 2008 (Walkup et al. 2008). Three main findings emerged. First, no model produced clearly significant results when using *R*^2^. Second, sMRI outperformed rsFC for fingerprinting, where it achieved excellent accuracy. Finally, both sMRI and rsFC methods had limitations; no single model emerged as consistently better than all other models, and prediction arose from hundreds of regions that did not cohere into networks identified in other studies.

An advantage of CPM over other predictive models concerns its capacity to generate interpretable findings that might prove useful in a clinical context. This contrasts with other machine-learning algorithms that are less widely used and that employ less discernible approaches with less clear applicability in such clinical contexts. Nevertheless, with CPM, the current findings suggest the need for improvements, before clinically useful approaches can emerge. For example, the edges that drove successful prediction varied across cross-validation loops. Such patterns complicate attempts to identify one set of robustly predictive edges. Findings in the current study also failed to reveal patterns closely overlapping with regions previously associated with anxiety. Of note, the lack of an established pattern does resemble the findings in another study linking resting-state networks to pediatric anxiety (Linke et al. 2021). While networks emerged in the prior study that differentiated anxiety from other forms of pediatric psychopathology, these networks also spanned multiple brain regions, generally not following networks derived from other studies.

As in Linke et al. (2021), the current findings could reflect a “many-to-one” pattern, where complex collections of connections in the brain interact to shape pediatric psychopathology. However, the lack of a recognizable network might also be related to methodological pitfalls of how the feature selection is done in CPM. The selection of features by using an arbitrary p-value threshold might result in the inclusion of edges that are not truly representative. It is often the case that in predictive work, there will be more neuroimaging features than subjects, making some form of feature selection necessary.

The prediction of therapeutic response in the current study went beyond a mere exercise of rating unseen data; it related functional connectivity to a gold-standard, clinically relevant outcome variable, over and above baseline levels of severity, as well as demographic factors such as age and sex. It is important to note that models that used imaging data to predict the post treatment PARS with baseline PARS as nuisance resulted in higher quality models than without. This is somewhat to be expected as the subject baseline symptom level might be related to brain measures and treatment outcomes.

Prediction offers potential to stratify subjects according to the likelihood that treatment is successful, to indicate those who may need additional support, as well as to use data to support mechanistic theories of psychopathology and their links to novel therapeutics, although some have warned caution (Mitchell et al. 2021). In effect, precision medicine and personalized clinical predictions have been garnering increased attention in recent years (Fair and Yeo 2020; Laumann, Zorumski, and Dosenbach 2023). However, a recent systematic review of 308 prediction models for psychiatry outcomes reported that 95% of studies were at high risk of bias primarily due to overfitting and biased variable selection methods and only 20% performed external validation on an independent sample (Meehan et al. 2022), highlighting the need to emphasize robust methodology and external validation in clinical models.

It is important to note that our study presents a relatively small sample size. The limited number of subjects included in each dataset led us to perform a leave-one-out cross-validation, which can yield unstable estimates of accuracy (Varoquaux et al. 2017). This is somewhat countered by the fact that we were able to include another dataset with similar inclusion criteria and study design as a validation set. Additionally, subjects underwent similar, but not identical treatment protocols in each dataset, as well as similar, but not identical imaging protocols. Finding replicable prediction despite such differences suggests the potential for improvements.

Neuroimaging studies including many sites are becoming more common. However, this is still not the case for neuroimaging studies embedded within clinical trials. In fact, the current study is the largest to use imaging data in a predictive framework in research on pediatric anxiety disorders. Extension to large multisite trials poses problems, since data on efficacy suggest that larger clinical trials for common pediatric mental illnesses contain more sources of noise than smaller studies with fewer study sites (Walkup 2017). This creates problems when attempting to maximize sample sizes through multi-site imaging studies embedded within clinical trials. Again, in the current study, replication in a new, albeit small, sample studied with somewhat different methods provides some evidence to generalizability (or the lack thereof).

As in a previous study, there was no overlap between the edges selected during CPM and the edges used for subject identification (Mantwill et al. 2022). However, the present data suggest that edge selection from fingerprinting using the most discriminatory features led to comparable results to the CPM. The edges selected in each case possibly represent different sources of variability that are not related; both approaches to edge selection might contain relevant, yet distinct information (Finn and Rosenberg 2021). However in our study no approach appeared particularly promising.

One factor limiting applicability concerned model selection: we successfully found models that appeared to be successful when using the correlation coefficient, but that are in fact predicting the target value worse than the mean. The *R*^2^ is a more adequate metric than the *r* value, and the results presented here highlights the need of using more than one metric when assessing predictive models.

Another limiting factor to the application of CPM in our sample is that the best models show a MAE of approximately 3.4 (PARS ranges between 0 and 25). Using predictive models in clinical practice is an emerging science. The added value that these models can bring to clinical practice remains uncertain and needs to be assessed objectively. In a study to detect the risk of mental health crisis stratified according to an automated model based on health registry, most clinical teams found the measure useful, although leading to relatively few additional actions (Garriga et al. 2022). Another topic that needs to be addressed is the need to train and test such predictive algorithms in more diverse settings. Our sample was mostly comprised of white Americans with high family income. Future studies should test if predictive models are generalizable to populations of diverse ethnic and cultural backgrounds.

Resting state fMRI is frequently criticized as an inaccurate picture of what would be brain resting-state activity, given that rsFC has been shown to relate to numerous uncontrolled variables such as mood (Harrison et al. 2008) or alertness (Chang et al. 2016), albeit more consistent results can be found using rsFC in predictive models than with other analytical approaches (Taxali et al. 2021). In effect, analysis of task-based fMRI from the same trial shows baseline differences and a return to normality after CBT (Haller et al. 2024). Additional use of tasks meant to draw out individual differences in the measure of interest may provide additional predictive power in CPM and reduce confounding effects due to the lack of engagement during rest (Finn et al. 2017). Greene and colleagues (2018), shows superior performance from task-based FC over resting state in predicting fluid intelligence, using multiple tasks, which varied in prediction accuracy by task. Rosenberg and colleagues (2016) uses both sustained attention task-based functional connectivity and resting state connectivity and finds significant predictions using both task and networks obtained from task with resting state data, though they do not compare accuracy of resting state CPM with task-based CPM. Barron and colleagues (2021) shows CPM can predict performance in memory tasks in a sample of participants with heterogeneous psychiatric disorders, a result which could be generalized to a healthy population. Using a general functional connectivity (GFC) measure based on multiple fMRI tasks may also have advantages over single-task CPM. Elliott and colleagues (2019) show increased test-retest reliability and higher heritability in GFC than rsFC. GFC may also improve prediction over single task FC, both when computed using averaged connectomes or concatenation of time series (Gao et al. 2019).

Our findings that resting state CPM can not reliably predict PARS score after treatment illustrate the difficulty in applying potential imaging-based measurements to improve treatment outcome predictions in a clinical sample. Additional research is necessary to explore how integrating different imaging modalities might benefit predictive algorithms. Our group is currently gathering fMRI data collected in pediatric patients seeking treatment for anxiety, collected during a naturalistic movie meant to induce anxiety and stronger individual differences that may relate to treatment response. The added value of movie-watching and task-based fMRI (Finn 2021) to predictive algorithms may help discover clinically relevant markers.

## Supporting information

Supplementary Results

## Data Availability

Data from some participants of the present study who may have consented to public data sharing are available upon request.

## Acknowledgements

This work was supported by the Intramural Research Program of the National Institutes of Health (NIH) through ZIA-MH002781 (https://clinicaltrials.gov: NCT00018057). AMW receives support from the NIH through U54-HG013247. This work used computational resources of the NIH Biowulf cluster (http://hpc.nih.gov).

## Footnotes

1 5 subjects in the final sample had TE = 3.42 ms and TI = 425 ms; the other parameters were the same.

