## Supplementary Material for "Brain Functional Connectivity and Anatomical Features as Predictors of Cognitive Behavioral Therapy Outcome for Anxiety in Youths"

Andre Zugman[1*], Grace V. Ringlein[1], Emily S. Finn[2], Krystal M. Lewis[1], Erin Berman[1],

Wendy K. Silverman[3], Eli R. Lebowitz[3], Daniel S. Pine[1] and Anderson M. Winkler[4].

1. Emotion and Development Branch, National Institute of Mental Health, National Institutes of Health, 9000 Rockville Pike, Bethesda, MD, 20892, USA.

2. Psychological and Brain Sciences, Dartmouth College, 3 Maynard St, Hanover, NH, 03755, USA.

3. Child Study Center, Yale University, 230 South Frontage Rd., New Haven, CT 06520, USA.

4. Division of Human Genetics, School of Medicine, University of Texas Rio Grande Valley, 1 West University Blvd, Brownsville, TX 78520, USA.

*Corresponding author: Emotion and Development Branch, National Institute of Mental Health, National Institutes of Health, 9000 Rockville Pike, Bethesda, MD, 20892, USA. Telephone: +1 (301) 480-8395,

### Supplementary Results

#### Processing choices

Across all measures, fingerprinting allowed identification of subjects based on their brain imaging profiles at rates that far exceeded chance. Nevertheless, the accuracy of fingerprinting, as well as that of CPM/APM, varied according to choices made during processing. Global signal regression (GSR) improved fingerprinting accuracy (53/66 vs 46/66 for baseline used to identify follow-up; 56/66 vs. 48/66 for follow-up used to identify baseline) as well as CPM accuracy (cross-validation: MAE difference = 0.0903, $p = 0.0003$; external validation: MAE difference = 0.1005, $p = 0.0001$). For these comparisons we used MAE as it uses the same units of the predicted quantities (PARS), thus serving as a better proxy of the size of the effect attributable to these differences. Using partial correlations instead of simple correlations reduced accuracy of fingerprinting (53/66 vs 33/66 for baseline used to identify follow-up; 56/66 vs. 32/66 for follow-up used to identify baseline), but improved accuracy of CPM (cross-validation: MAE difference = 0.2101, $p = 0.0001$; external validation: MAE difference = 0.1154, $p = 0.0002$). However, other processing choices also affected accuracy, and the generally better models used full (not partial) correlations.

Likewise, the accuracy of APM and anatomical fingerprinting varied depending on processing choices, although with a less consistent pattern than for CPM. Accuracy was perfect or near perfect for area, thickness, curvature, and sulcal depth, with at least 64/66 correct identifications for all these measurements, with or without smoothing, but not as close to perfect for gray/white matter contrast (59/66 with smoothing, although 64/66 without). Gray/white matter contrast, however, generally produced the best APM results compared, for example, with thickness (cross-validation: MAE difference = 6.1119, $p = 0.0001$; external validation: MAE difference =

0.2528, p = 0.0001) or surface area (cross-validation: MAE difference = 1.2642, p = 0.0100; external validation: MAE difference = 0.1274, p = 0.0001). While smoothing reduced the accuracy of fingerprinting only for gray/white matter contrast without affecting the accuracy for other anatomical measures, it did, counterintuitively, reduce the accuracy of APM for most models and measurements considered. For gray/white matter contrast, smoothing in some cases led to no vertices being detected in the first stage of the predictive modeling.

For both CPM and APM, using (a) a regression model that did not include age, sex, and scanner (where applicable) as nuisance variables, (b) a model in which these were regressed out from data and design, or (c) a model in which these were used as predictors, led to sometimes improved or reduced MAE. Likewise, the choice of edges in the first stage of CPM as (a) positively correlated only, (b) negatively correlated only, (c) both, or (d) the edges with highest differential power from fingerprinting, also led to inconsistent improvements or reductions in the MAE. Weighting the edges by their p-value in logarithmic scale (a rough measure of effect size given the fixed sample sizes) also sometimes improved, sometimes reduced accuracy. The Supplementary Material to this paper includes a large spreadsheet file containing accuracy results (MAE, r, and R2) for CPM and APM, in both cases using cross-validation within Dataset A and external validation using Dataset B. In the spreadsheet, the columns represent:

- *filename:* A unique identifier for the text file containing the results shown in the corresponding row (not relevant for reading these results).
- *model:* Indicates whether the model included variables such as age, sex, and baseline PARS were used as predictors (predictors), as nuisance variables (residualized), or if omitted from the model (nonuisance).

- *meas (APM only):* Indicates what morphological variable was used for prediction: area (area), curvature (curv), sulcal depth (sulc), cortical thickness (thickness), or gray/white contrast (w-g.pct.mgh).
- *fwhm (APM only):* Indicates the amount of smoothing applied: no smoothing (0) or smoothing with a Gaussian kernel of full width at half maximum of 15 mm (15).
- *denoise (CPM only):* Indicates whether denoising used AROMA components and white matter and CSF signal (AROMA) or further included global signal regression (AROMA-GSR).
- *netmat (CPM only):* Indicates whether the connectivity matrix used for this model used partial correlations (partial) or not (full).
- *vertices (APM) or edges (CPM):* Indicates whether edges/vertices selected in the first regression of the predictive modeling were those with a positive (pos) correlation with PARS, negative (neg), or both (both), or if the edges/vertices selected were those with highest differential power from fingerprinting (finger).
- *weighted:* Indicates whether a simple sum of values from edges/vertices selected in the first stage of the predictive model was used (FALSE) or whether a weighted sum based on the negative logarithm of p-values was used (TRUE), thus giving stronger weight to more significant edges/vertices.
- *pars:* Indicates what PARS was tentatively predicted by the model: PARS at the start of treatment (i.e., week 0, parsTotalStart), PARS at the end of treatment (i.e., week 8 or 12 depending on the dataset, parsTotalEnd), the difference between PARS at start and end (parsDelta), or PARS at the end of treatment after taking the PARS at start as nuisance (BaselineAsNuisance). For BaselineAsNuisance, there are no models configured as nonuisance.
- *train, test (external validation only):* Indicators of what datasets were used for training or for testing (i.e., datasets A or B). The spreadsheet also includes training and testing on

the same dataset; such models are expected to have excellent performance, and were run only for sanity checking; results are not meant to be used otherwise.

- *Rsq, Rsq_lowerCI, Rsq_upperCI:* Cross-validation R2, along with lower and upper bounds for the corresponding 95% bootstrap confidence intervals.
- *MAE, MAE_lowerCI, MAE_upperCI:* Mean absolute error, along with lower and upper bounds for the corresponding 95% bootstrap confidence intervals.
- *Corr, Corr_lowerCI, Corr_upperCI:* Correlation between observed and predicted values, along with lower and upper bounds for the corresponding 95% bootstrap confidence intervals.
- *NumberBootStraps:* Number of bootstraps used to produce the confidence intervals.
- *NumberOfVertices (APM) or NumberOfEdges (CPM):* For external validation, this is the number of vertices or edges selected in the first stage of the predictive modeling. For cross-validation, this is the average across leave-one-out folds. If there are zero edges or vertices selected in the first stage, models configured as predictors and residualized are expected to yield the same results.

Colors in the spreadsheet are conditional on the values shown, and convey no information other than already represented by the respective numbers.

### Supplementary Tables and Figures

**Supplementary Table 1: Detailed descriptive statistics for Datasets A and B, as used for the CPM and APM analyses.**

| | Dataset A (n = 54) | | Dataset B (n = 15) | | | |
|---|---|---|---|---|---|---|
| | Mean or n | SD or % | Mean or n | SD or % | Test statistic* | p-value |
| Age | 13.06 | 2.70 | 13.19 | 3.37 | -0.17 | 0.87 |
| Framewise displacement (mean) | 0.11 | 0.05 | 0.14 | 0.06 | -2.15 | 0.03 |
| Framewise displacement (%) | 9.79 | 8.88 | 15.40 | 13.34 | -1.93 | 0.06 |
| IQ (WASI) | 112.39 | 13.42 | 115.53 | 17.17 | -0.75 | 0.45 |
| PARS total at baseline | 14.63 | 2.84 | 16.73 | 2.89 | -2.53 | 0.01 |
| PARS total at follow-up | 8.93 | 4.44 | 13.73 | 3.37 | -3.88 | <0.01 |
| PARS difference | -5.70 | 4.39 | -3.00 | 3.98 | -2.15 | 0.04 |
| Sex: | | | | | 0.01 | 0.93 |
| Female | 33 | 61 % | 10 | 67 % | | |
| Male | 21 | 39 % | 5 | 33 % | | |
| Race (as reported): | | | | | 6.80 | 0.34 |
| American Indian or Alaskan Native | 1 | 2 % | 0 | | | |
| Asian | 2 | 4 % | 1 | 7 % | | |
| Black or African American | 2 | 4 % | 2 | 13 % | | |
| Multiple Races | 8 | 15 % | 1 | 7 % | | |
| Unknown | 3 | 6 % | 1 | 7 % | | |
| White | 38 | 70 % | 9 | 60 % | | |
| Ethnicity: | | | | | 1.03 | 0.60 |
| Latino or Hispanic | 9 | 17 % | 2 | 13 % | | |
| Not Latino or Hispanic | 42 | 78 % | 13 | 87 % | | |
| Unknown | 3 | 6 % | 0 | | | |
| Highest Educational level of the parents: | | | | | 3.34 | 0.50 |
| Graduate Level | 41 | 76 % | 10 | 67 % | | |
| Standar College | 8 | 15 % | 5 | 33 % | | |
| Partial College, High School or lower | 4 | 8 % | 0 | | | |
| Income: | | | | | 9.39 | 0.05 |
| < $60,000 | 0 | 0% | 2 | 13% | | |
| $60,000.00 - $89,999.99 | 6 | 11 % | 4 | 27 % | | |
| $90,000.00 - $179,999.99 | 20 | 37 % | 4 | 27 % | | |
| > $180,000.00 | 24 | 44 % | 5 | 33 % | | |
| Diagnosis**: | | | | | 7.50 | 0.76 |
| Panic Disorder | 2.0 | 4 % | 0.0 | 0 % | | |
| Separation Anxiety | 17.0 | 31 % | 5.0 | 33 % | | |
| Social Phobia | 33.0 | 61 % | 10.0 | 67 % | | |
| Specific Phobia | 14.0 | 26 % | 3.0 | 20 % | | |
| Generalized Anxiety Disorder | 46.0 | 85 % | 11.0 | 73 % | | |

* Two-sample t-test or Chi-squared test when appropriate.

** Patients may have more than one diagnosis, thus the sum is higher than 100%.

**Supplementary Table 2: Comparison between included and excluded subjects in the CPM and APM analyses – Dataset A.**

| | Included (n = 54) | | Excluded (n = 12) | | | |
|---|---|---|---|---|---|---|
| | Mean or n | SD or % | Mean or n | SD or % | Test statistic* | p-value |
| Age | 13.06 | 2.70 | 12.17 | 3.26 | 0.99 | 0.32 |
| Framewise displacement (mean) | 0.11 | 0.05 | 0.70 | 0.83 | -5.32 | 0.00 |
| Framewise displacement (%) | 9.79 | 8.88 | 46.99 | 19.50 | -10.20 | 0.00 |
| IQ (WASI) | 112.39 | 13.42 | 118.33 | 11.53 | -1.42 | 0.16 |
| PARS total at baseline | 14.63 | 2.84 | 15.83 | 3.19 | -1.30 | 0.20 |
| PARS total at follow-up | 8.93 | 4.44 | 9.17 | 5.24 | -0.16 | 0.87 |
| PARS difference | -5.70 | 4.39 | -6.67 | 4.01 | 0.70 | 0.49 |
| Sex: | | | | | 0.15 | 0.70 |
| Female | 33 | 61 % | 6 | 50 % | | |
| Male | 21 | 39 % | 6 | 50 % | | |
| Race (as reported): | | | | | 2.42 | 0.79 |
| American Indian or Alaskan Native | 1 | 2 % | 0 | | | |
| Asian | 2 | 4 % | 0 | | | |
| Black or African American | 2 | 4 % | 0 | | | |
| Multiple Races | 8 | 15 % | 3 | 25 % | | |
| Unknown | 3 | 6 % | 0 | | | |
| White | 38 | 70 % | 9 | 75 % | | |
| Ethnicity: | | | | | 1.05 | 0.59 |
| Latino or Hispanic | 9 | 17 % | 3 | 25 % | | |
| Not Latino or Hispanic | 42 | 78 % | 9 | 75 % | | |
| Unknown | 3 | 6 % | 0 | | | |
| Highest educational level of the parents: | | | | | 0.47 | 0.79 |
| Graduate Level | 41 | 76 % | 9 | 75 % | | |
| Standar College | 8 | 15 % | 2 | 17 % | | |
| Partial College, High School or lower | 2 | 4 % | 0 | 0 % | | |
| Income: | | | | | 1.40 | 0.50 |
| < $60,000 | 0 | 0 % | 0 | 0 % | | |
| $60,000.00 - $89,999.99 | 6 | 11 % | 1 | 8 % | | |
| $90,000.00 - $179,999.99 | 20 | 37 % | 6 | 50 % | | |
| > $180,000.00 | 24 | 44 % | 3 | 25 % | | |
| Diagnosis**: | | | | | 15.21 | 0.23 |
| Panic Disorder | 2.0 | 4 % | 1.0 | 8 % | | |
| Separation Anxiety | 17.0 | 31 % | 4.0 | 33 % | | |
| Social Phobia | 33.0 | 61 % | 7.0 | 58 % | | |
| Specific Phobia | 14.0 | 26 % | 5.0 | 42 % | | |
| Generalized Anxiety Disorder | 46.0 | 85 % | 10.0 | 83 % | | |

* Two-sample t-test or Chi-squared test when appropriate.

** Patients may have more than one diagnosis, thus the sum is higher than 100%.

**Supplementary Table 3: Comparison between included and excluded subjects in the CPM and APM analyses – Dataset B.**

| | Included (n = 15) | | Excluded (n = 16) | | | |
|---|---|---|---|---|---|---|
| | Mean or n | SD or % | Mean or n | SD or % | Test statistic* | p-value |
| Age | 13.19 | 3.37 | 10.33 | 1.24 | 3.18 | 0.00 |
| Framewise displacement (mean) | 0.14 | 0.06 | 0.77 | 0.58 | -4.15 | 0.00 |
| Framewise displacement (%) | 15.40 | 13.34 | 54.24 | 16.55 | -7.16 | 0.00 |
| IQ (WASI) | 115.53 | 17.17 | 109.00 | 18.04 | 1.02 | 0.32 |
| PARS total at baseline | 16.73 | 2.89 | 16.44 | 3.05 | 0.28 | 0.78 |
| PARS total at follow-up | 13.73 | 3.37 | 12.69 | 3.46 | 0.85 | 0.40 |
| PARS difference | -3.00 | 3.98 | -3.75 | 3.53 | 0.56 | 0.58 |
| Sex: | | | | | 0.02 | 0.89 |
| Female | 10 | 67 % | 10 | 62 % | | |
| Male | 5 | 33 % | 6 | 38 % | | |
| Race (as reported): | | | | | 2.50 | 0.78 |
| American Indian or Alaskan Native | 1 | 7 % | 0 | | | |
| Asian | 2 | 13 % | 2 | 12 % | | |
| Black or African American | 1 | 7 % | 2 | 12 % | | |
| Multiple Races | 1 | 7 % | 0 | | | |
| Unknown | 1 | 7 % | 1 | 6 % | | |
| White | 9 | 60 % | 11 | 69 % | | |
| Ethnicity: | | | | | 4.01 | 0.13 |
| Latino or Hispanic | 2 | 13 % | 0 | | | |
| Not Latino or Hispanic | 13 | 87 % | 14 | 88 % | | |
| Highest educational level of the parents: | | | | | 0.00 | 1.00 |
| Graduate Level | 10 | 67 % | 11 | 69 % | | |
| Standar College | 5 | 33 % | 4 | 25 % | | |
| Partial College, High School or lower | 0 | 0 % | 0 | 0 % | | |
| Income: | | | | | 5.90 | 0.12 |
| < $60,000 | 2 | 13 % | 2 | 12 % | | |
| $60,000.00 - $89,999.99 | 4 | 27 % | 0 | 0 % | | |
| $90,000.00 - $179,999.99 | 4 | 27 % | 9 | 56 % | | |
| > $180,000.00 | 5 | 33 % | 5 | 31 % | | |
| Diagnosis**: | | | | | 8.04 | 0.53 |
| Panic Disorder | 0.0 | 0 % | 0.0 | 0 % | | |
| Separation Anxiety | 5.0 | 33 % | 9.0 | 56 % | | |
| Social Phobia | 10.0 | 67 % | 8.0 | 50 % | | |
| Specific Phobia | 3.0 | 20 % | 4.0 | 25 % | | |

* Two-sample t-test or Chi-squared test when appropriate.

** Patients may have more than one diagnosis, thus the sum is higher than 100%.

**Supplementary Table 4 : Comparison between included and excluded subjects in the fingerprinting analyses.**

| | Included (n = 66) | | Excluded (n = 21) | | | |
|---|---|---|---|---|---|---|
| | Mean or n | SD or % | Mean or n | SD or % | Test statistic* | p-value |
| Age at baseline | 13.86 | 2.59 | 12.29 | 2.93 | 2.30 | 0.02 |
| Age at follow-up | 14.14 | 2.56 | 12.68 | 2.97 | 2.14 | 0.03 |
| Time between scans (days) | 122.78 | 40.37 | 142.76 | 55.55 | -1.79 | 0.08 |
| IQ | 111.52 | 12.83 | 116.37 | 11.49 | -1.49 | 0.14 |
| Sex: | | | | | 0.21 | 0.64 |
| Female | 39 | 59 % | 10 | 48 % | | |
| Male | 27 | 41 % | 10 | 48 % | | |
| Race (as reported): | | | | | 2.25 | 0.81 |
| American Indian or Alaskan Native | 1 | 2 % | 0 | | | |
| Asian | 4 | 6 % | 1 | 5 % | | |
| Black or African American | 6 | 9 % | 1 | 5 % | | |
| Multiple Races | 13 | 20 % | 4 | 19 % | | |
| Unknown | 4 | 6 % | 0 | | | |
| White | 38 | 58 % | 14 | 67 % | | |
| Ethnicity: | | | | | 0.71 | 0.70 |
| Latino or Hispanic | 8 | 12 % | 3 | 14 % | | |
| Not Latino or Hispanic | 56 | 85 % | 17 | 81 % | | |
| Unknown | 2 | 3 % | 0 | | | |
| Highest educational level of the parents: | | | | | 1.87 | 0.39 |
| Graduate Level | 42 | 64 % | 13 | 62 % | | |
| Standar College | 12 | 18 % | 5 | 24 % | | |
| Partial College, High School or lower | 5 | 8 % | 0 | 0 % | | |
| Any psychiatric diagnosis: | | | | | 1.20 | 0.27 |
| Yes | 42 | 64 % | 16 | 76 % | | |
| No | 24 | 36 % | 4 | 19 % | | |
| Income: | | | | | 5.78 | 0.12 |
| < $60,000.00 | 5 | 8 % | 0 | 0 % | | |
| $60,000.00 - $89,999.99 | 5 | 8 % | 3 | 14 % | | |
| $90,000.00 - $179,999.99 | 21 | 32 % | 10 | 48 % | | |
| > $180,000.00 | 27 | 41 % | 4 | 19 % | | |

* Two-sample t-test or Chi-squared test when appropriate.

**Supplementary Table 5: Detailed descriptive statistics for the fingerprinting sample. In the fingerprinting analysis HV were included to maximize the sample with baseline and follow-up scans.**

| | Patients (n = 42) | | HV (n = 24) | | | |
|---|---|---|---|---|---|---|
| | Mean or n | SD or % | Mean or n | SD or % | Test statistic * | p-value |
| Time between scans (days) | 128.20 | 40.68 | 113.54 | 38.93 | 1.42 | 0.16 |
| Age at baseline | 13.67 | 2.67 | 14.19 | 2.47 | -0.78 | 0.44 |
| Age at follow-up | 13.93 | 2.63 | 14.50 | 2.45 | -0.86 | 0.39 |
| FD (mean) at baseline | 0.11 | 0.05 | 0.10 | 0.05 | 0.17 | 0.87 |
| FD (%) at baseline | 9.20 | 9.23 | 9.88 | 11.86 | -0.26 | 0.79 |
| FD (mean) at follow-up | 0.12 | 0.05 | 0.11 | 0.05 | 0.49 | 0.62 |
| FD (%) at follow-up | 10.21 | 9.51 | 10.24 | 10.07 | -0.01 | 0.99 |
| IQ (WASI) | 111.33 | 13.06 | 111.83 | 12.67 | -0.15 | 0.88 |
| Sex: | | | | | 0.77 | 0.38 |
| Female | 27 | 64 % | 12 | 50 % | | |
| Male | 15 | 36 % | 12 | 50 % | | |
| Race (as reported): | | | | | 7.76 | 0.17 |
| American Indian or Alaskan Native | 1 | 2 % | 0 | | | |
| Asian | 2 | 5 % | 2 | 8 % | | |
| Black or African American | 1 | 2 % | 5 | 21 % | | |
| Multiple Races | 8 | 19 % | 5 | 21 % | | |
| Unknown | 3 | 7 % | 1 | 4 % | | |
| White | 27 | 64 % | 11 | 46 % | | |
| Ethnicity: | | | | | 1.80 | 0.41 |
| Latino or Hispanic | 6 | 14 % | 2 | 8 % | | |
| Not Latino or Hispanic | 34 | 81 % | 22 | 92 % | | |
| Unknown | 2 | 5 % | 0 | | | |
| Highest educational level of the parents: | | | | | 6.95 | 0.03 |
| Graduate Level | 34 | 81 % | 8 | 33 % | | |
| Standar College | 6 | 14 % | 6 | 25 % | | |
| Partial College, High School or lower | 2 | 5 % | 3 | 12 % | | |
| Income: | | | | | 7.07 | 0.07 |
| < $60,000 | 1 | 2 % | 4 | 17 % | | |
| $60,000.00 - $89,999.99 | 4 | 10 % | 1 | 4 % | | |
| $90,000.00 - $179,999.99 | 13 | 31 % | 8 | 33 % | | |
| > $180,000.00 | 21 | 50 % | 6 | 25 % | | |
| Diagnosis**: | | | | | | |
| Panic Disorder | 2 % | 5 % | | | | |
| Separation Anxiety | 11 % | 26 % | | | | |
| Social Phobia | 27 % | 64 % | | | | |
| Specific Phobia | 13 % | 31 % | | | | |
| Generalized Anxiety Disorder | 36 % | 86 % | | | | |

* Two-sample t-test or Chi-squared test when appropriate.

** Patients may have more than one diagnosis, thus the sum is higher than 100%.

**Supplementary Figure 1: Pairplot graph of DP (differential power) by modality.**

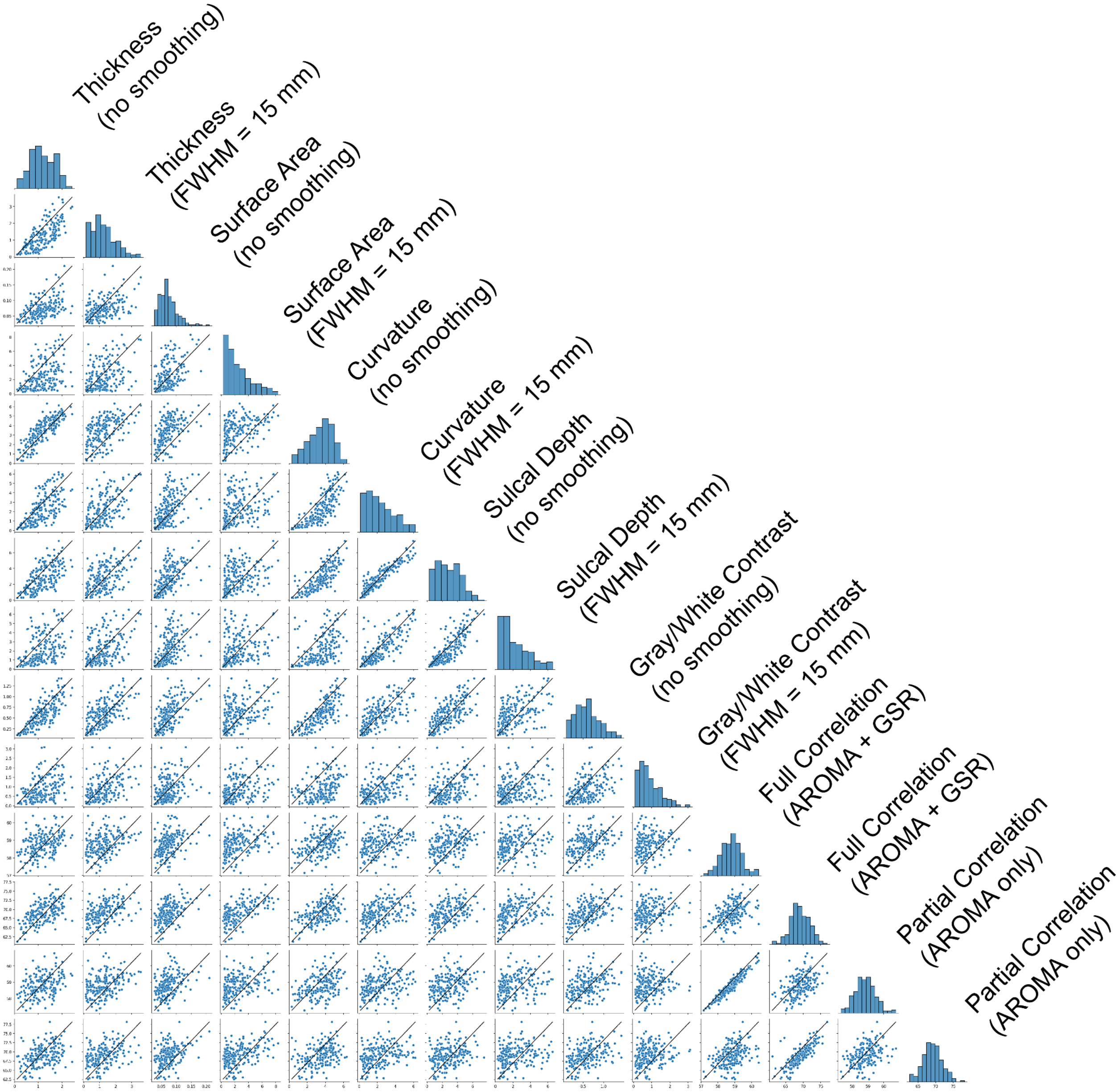


Pairplot graph of DP (differential power) by modality, including anatomical measurements (thickness, surface area, curvature, sulcal depth, and gray/white contrast) and functional measurements (connectivity assessed with full or partial correlations). For anatomical measurements, results for two levels of smoothing are shown; for functional, results are shown for denoising with or without global signal regression (GSR). Scatterplots represent the DP in 200 cortical ROIs in each modality. For vertex-wise data, the DP for all vertices within the ROI were averaged. FC is the average value of the average DP for all edges that represents connection to the ROI. The diagonal displays the histogram for the ROI DP in each modality.
